# Environmental predictors of malaria infection in Sussundenga, Mozambique

**DOI:** 10.1101/2023.06.29.23292060

**Authors:** Alexa Steiber, João L. Ferrão, Albino B. Francisco, Valy Muhiro, Anísio Novela, Dominique E. Earland, Kelly M. Searle

**Author notes:** Corresponding Author: Alexa Steiber.

## Abstract

Malaria is highly sensitive to environmental conditions, including climate variability and land use practices. Ecologically, Sussundenga district has a significantly lower elevation compared to the Zimbabwe border and a more tropical climate compared to southern and northern Mozambique due to high seasonal rainfall. We aimed to evaluate the effects of climate and environmental factors at the household level on rapid diagnostic test results for Malaria in Sussundenga, Mozambique. To understand this association, we collected publicly available United States Geological Survey satellite data on elevation, vegetation, and land use cover. Additionally, we collected satellite data on day and night land surface temperatures and evapotranspiration which we assessed at 1- and 2-week lags. We spatially and temporally joined these data with malaria infection data at the household level. Using this database, we assessed whether these environmental factors were good predictors for having a positive rapid diagnostic test result using spatio-temporal models that accounted for the underlying correlation structure. Risk factor surveillance is an important tool for controlling the spread of infectious diseases. The results from modeling of the ecological predictors of malaria infection and spatial maps provided in this study could aid in developing frameworks to mitigate malaria transmission and predict future malaria transmission in this region. Understanding how environmental changes impacts malaria transmission and infection at the household level may have important implications for vector control and disease surveillance strategies utilized by the district.

## Introduction

Malaria is a life-threatening disease that is transmitted to humans through the bite of *Anopheles spp*. mosquitoes infected with *Plasmodium spp*. parasites. In 2021, Mozambique had a cumulative incidence of 32% and was among the four countries that accounted for almost half of all malaria cases globally (the other three being Nigeria, Democratic Republic of the Congo, and Uganda)(1). Malaria is endemic in Mozambique, with seasonal peaks ranging from December through April depending on the ecological zone and length of the rainy season.

Malaria transmission is influenced by numerous environmental and ecological conditions. Variation in climatic conditions, such as temperature, rainfall patterns, and humidity, have a profound effect on the longevity and fecundity of the mosquito vectors and on the development of *Plasmodium falciparum* parasites in the mosquito vectors, which is why increases in malaria incidence, prevalence, and distribution have been identified as potential consequences of climate change(2). The Notre Dame Global Adaptation Initiative (ND-GAIN) summarizes countries’ vulnerability to climate change by assessing their economic, social, and governance readiness, their potential for exposure to climate change, the degree to which they depend on sectors that are impacted by climate change, and their adaptive capacity(3). In 2020, based on the aforementioned factors, ND-GAIN ranked Mozambique as the 48^th^ out of 182 most vulnerable to, and the 21^st^ out of 192 least ready to address the effects of climate change(3).

Despite the burden of malaria in Mozambique, and its vulnerability to climate change, the use of spatio-temporal models to examine the association between malaria risk and environmental factors has been relatively underutilized. One study conducted in Sussundenga Village, Mozambique which aimed to develop malaria risk maps as a function of environmental conditions found that precipitation, slope, and land cover were related to malaria risk.(4) In addition to environmental variables, a systematic review conducted in 2015 found that improved housing features are associated with a lower risk of epidemiological outcomes, including malaria infection.(5) Manica Province in central Mozambique has the second highest malaria incidence in the country and Sussundenga village is one of the most affected areas in this province.^4^ The objective of this study was to integrate individual level, household level, and environmental level data, across space and time, in order to evaluate the impacts of environmental factors and housing structure on malaria infection measured by rapid diagnostic test (RDT) in Sussundenga Village.

## Materials and Methods

### Study area

Sussundenga village is an agrarian community located in Sussundenga District, Manica Province, Mozambique. It is a rural area located 42 km from Chimoio, the capital of Manica Province, and is approximately 40km from the Zimbabwe border.

**Figure 1:**
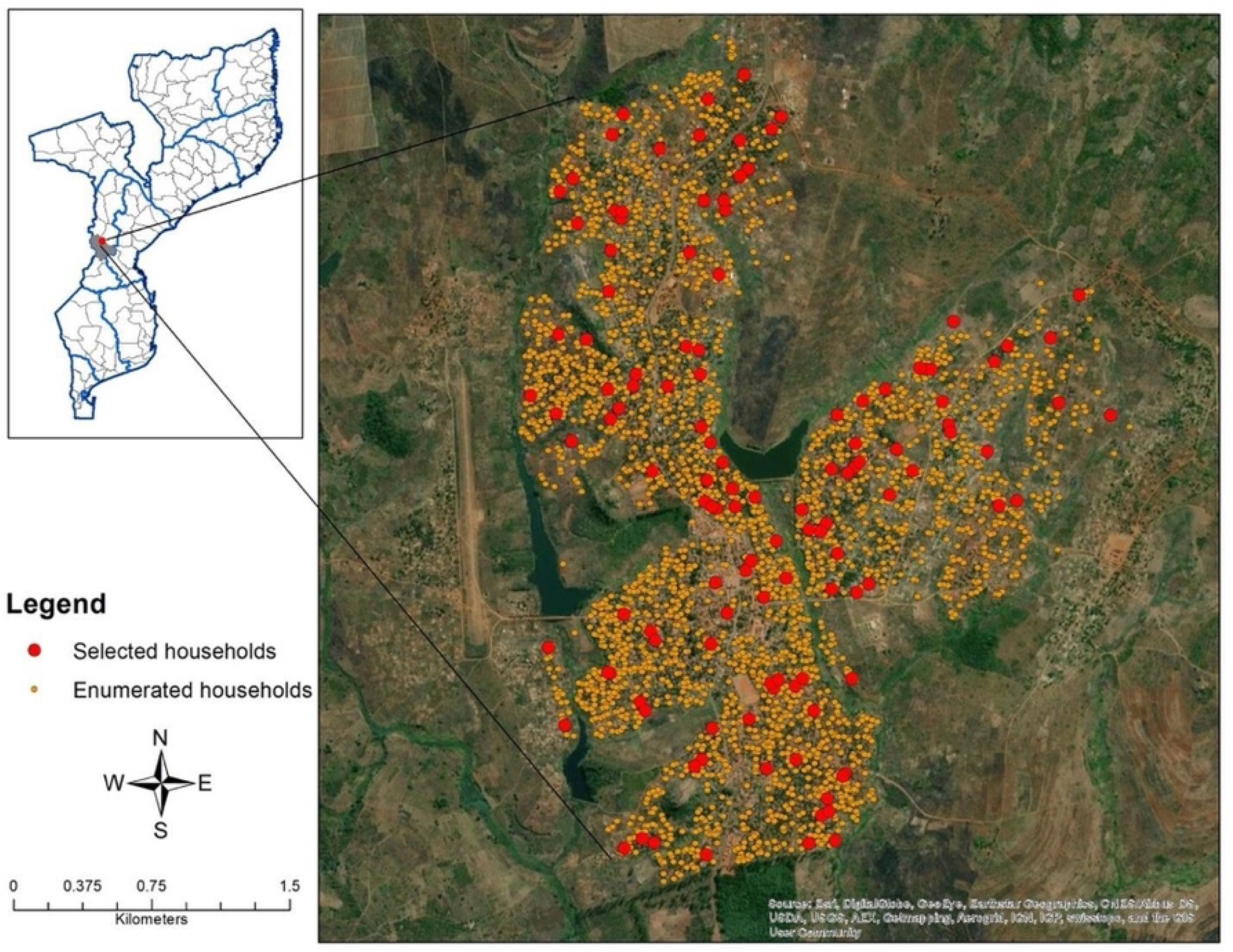
Study area in Sussundenga District, Mozambique.

Sussundenga has an estimated population of 31,429 individuals. Males and females comprise 47% and 53% of the population, respectively, and the majority of individuals are 15 years or older (50.6%).^4^

Ecologically, Sussundenga District has a significantly lower elevation compared to the Zimbabwe border and a more tropical climate compared to southern and northern Mozambique due to high seasonal rainfall.^5,6^) The rainy season occurs from November to April. The annual precipitation is 1200mm and the average temperature is 21.2 °C.^5^

### Data collection

Publicly available climate data was collected from the United States Geological Survey’s (USGS) Moderate Resolution Imaging Spectroradiometer (MODIS) and the NASA Digital Elevation Model (NASADEM) on day and night land surface temperature, evapotranspiration, normalized difference vegetation index (NDVI), land cover, and elevation.

#### Land Surface Temperature (LST)

The MOD11A2 Version 6 product provides an 8-day per-pixel Land Surface Temperature (day and night) with a 1-kilometer spatial resolution in a 1200 by 1200 km grid. Each pixel value in the MOD11A2 is a simple average of all the corresponding MOD11A2 LST pixels collected within that 8-day period.

#### Evapotranspiration (ET)

The MOD16A2 Version 6 ET product is an 8-day composite dataset produced at 500-meter pixel resolution. The algorithm used for the MOD16 data product collection is based on the logic of the Penman-Monteith equation, which includes inputs of daily meteorological reanalysis data along with MODIS remotely sensed data products such as vegetation property dynamics, albedo, and land cover (USGS MOD16A2 v006).(7) The pixel values for the ET layer is the sum of all eight days within the composite period.

#### Vegetation (NDVI)

The MOD13A1 Version 6 product provides NDVI values at a per pixel basis at 500-meter spatial resolution. MODIS vegetation indices, produced on 16-day intervals, provide consistent spatial and temporal comparisons of vegetation canopy greenness, a composite property of leaf area, chlorophyll and canopy structure.

#### Land Cover (LC)

The MCD12Q1 Version 6 data product provides global land cover types at yearly intervals, derived from six different classification schemes. The classifications are derived through a supervised decision-tree classification method and undergo additional post-processing that incorporate prior knowledge and ancillary information to further refine specific classes. The primary land cover scheme identifies 17 classes defined by the International Geosphere-Biosphere Programme (IGBP), including 11 natural vegetation classes, three human-altered classes, and three non-vegetated classes.

#### Elevation

Elevation data were obtained from NASADEM Version 1 dataset housed on the USGS Earth Explorer platform. NASADEM provides global elevation data at 1 arc second spacing.

Epidemiological data on individual characteristics and malaria infection status was obtained from a cross-sectional community survey conducted in Sussundenga village from December 2019-February 2020. Households were randomly selected for screening from satellite imagery after which data collectors used GPS coordinates to approach the selected households for participation. The data collectors enrolled 100 households, recorded the household GPS coordinates, and administered an electronic survey to all eligible participants (any full-time resident older than 3 months with parents providing responses for children aged 3 months to 12 years). Individual and household-level demographic information and housing structure information was collected via the electronic survey. Additionally, a study nurse collected a finger prick blood sample and administered an RDT [Right Sign Malaria Pf (Biotest, Hangzhou Biotest Biotech Co, China]. All data were collected and stored using the REDCap® server hosted at University of Minnesota School of Public Health and was treated confidentially.(8)

### Data analysis

All data analyses were performed using Stata (StataCorp LLC, version 16.1). All data were deidentified prior to analysis. The environmental and epidemiological data were spatially and temporally joined at the household level using QGIS (Matera, version 7.8.0). A binary variable was used to represent the dependent variable, malaria infection by RDT. Pearson Chi square tests for categorical variables and t-tests for independent variables were used to compare the following socio-demographic characteristics between RDT positive and negative individuals without adjusting for covariates:

- Individual level: Age, sex, ITN use the previous night
- Household level: Head of household occupation, head of household type of employment (full-time, part-time, seasonal), head of household highest education level, wall, floor, and roof material, whether the house had holes in the walls, whether the eaves were open, whether the windows closed (completely, partially, or not at all), and the vector control measure used (drains water around the house, cuts the grass/keeps the lawn clean, burns oils or mosquito coils, uses insecticides outside of the house, or none)
- Environment level: Landcover, elevation, ET, day LST, night LST, and NDVI

Univariate analyses on all explanatory variables were conducted. Day and night LSTs and ET were assessed at both 1- and 2-week lags. Only one of the time lagged variables for each factor (day and night LST, and ET)-the one that appeared to be the best predictor for malaria based on the effect size and statistical significance-was included in the final models. Interactions between the environmental predictors were assessed, however none were statistically significant and thus were not included in the final models. Forward selection was used to build the final generalized estimating equation (GEE) models that account for the underlying correlation structure (independent) in order to examine the association between each environmental factor and having a positive RDT. Household structural factors were assessed as effect modifiers in each of these models.

### Ethics Statement

Ethical review and approval for this study was completed by the Institutional Review Board (IRB) at the University of Minnesota [STUDY00007184] and from A Comissão Nacional de Bioética em Saúde (CNBS) at the Ministry of Health of Mozambique [IRB00002657]. Data collectors obtained verbal informed consent along with a written signature or thumb print (for record keeping purposes) for all adult residents and parent/guardian permission for children between 3 months and 12 years old and assent for minors between 13 and 17 years old.

## Results

### Demographic Characteristics

Of the 303 individuals tested, 94 (31%) were found to be positive for malaria by RDT at the time of the survey (Table 1). The median age of participants that tested positive was 12 and the median age of participants that tested negative was 20. Based on the 95%CIs, there was not a statistically significant difference regarding the proportion of males and females across malaria status groups; among those who tested positive, 46.7% were male and 53.3% were female and among those who tested negative, 43.9% were male and 56.1% were female. There was also not a statistically significant difference between those who had a positive RDT and a negative RDT in regard to whether they slept under an ITN the previous night.

**Table 1:** Demographic characteristics of participants

| Characteristic | RDT POS<br>(Just report<br>medians<br>and<br>percentages<br>) | IQR/CI | RDT NEG<br>(Just report<br>medians<br>and<br>percentage<br>s) | IQR/CI |
| --- | --- | --- | --- | --- |
|  | *n* = 94 |  | *n* = 209 |  |
| **Individual Level** |  |  |  |  |
| Age, years, median | 12 | 7-19 | 20 | 11-31 |
| Male gender, % | 46.7 | 0.36-0.57 | 43.9 | 0.37-0.51 |
| Female gender, % | 53.3 | 0.43-0.64 | 56.1 | 0.46-0.67 |
| Slept under ITN previous night, *n* (%) | 55.3 | 0.45-0.66 | 68.3 | 0.62-0.75 |
| **Household Level** |  |  |  |  |
| Head of household occupation, *n* (%) |  |  |  |  |
| *Farmer* | 52.1 | 0.42-0.63 | 38 | 0.31-0.45 |
| *Teacher* | 11.7 | 0.06-0.20 | 15.9 | 0.11-0.22 |
| *Healthcare worker* | 3.2 | 0.01-0.09 | 6.7 | 0.04-0.11 |
| *Shopkeeper* | 0 | 0-0.04 | 1.9 | 0.01-0.05 |
| *Other* | 33 | 0.24-0.43 | 37.5 | 0.31-0.44 |
| Head of household type of employment, *n* (%) |  |  |  |  |
| *Full time* | 48.9 | 0.39-0.60 | 69.9 | 0.63-0.76 |
| *Seasonally or Part time* | 51.1 | 0.41-0.62 | 30.1 | 0.24-0.37 |
| Head of household highest education level, *n* (%) |  |  |  |  |
| *Primary school 1 and 2* | 41.5 | 0.31-0.52 | 29.6 | 0.24-0.36 |
| *Secondary school 1* | 20.2 | 0.13-0.30 | 24.9 | 0.20-0.31 |
| *Technical school or university* | 25.6 | 0.17-0.36 | 35.4 | 0.29-0.42 |
| *No school* | 10.6 | 0.05-0.19 | 10.1 | 0.06-0.15 |
| *Do not know* | 2.1 | 0.003-0.08 | 0 | 0-0.02 |
| Wall material, *n* (%) |  |  |  |  |
| *Natural (i.e., no walls, straw)* | 4.3 | 0.01-0.11 | 7.3 | 0.04-0.12 |
| *Rudimentary (i.e., bamboo, zinc sheet)* | 41.4 | 0.31-0.52 | 27.2 | 0.21-0.34 |
| *Modern (i.e., cement blocks, bricks)* | 54.3 | 0.44-0.65 | 65.5 | 0.59-0.72 |
| House has holes in the wall, *n* (%) | 46.2 | 0.35-0.56 | 34 | 0.28-0.41 |
| Floor material, *n* (%) |  |  |  |  |
| *Natural (i.e., ground, clay floor)* | 58.5 | 0.48-0.69 | 38.3 | 0.32-0.45 |
| *Rudimentary (i.e., wood)* | 0 | 0-0.04 | 1.4 | 0.003-0.04 |
| *Modern (i.e., cement, brick, tile)* | 41.5 | 0.31-0.52 | 60.3 | 0.53-0.67 |
| Roof material, *n* (%) |  |  |  |  |
| *Natural (i.e., grass, leaves, or no roof)* | 8.5 | 0.04-0.16 | 4.8 | 0.02-0.09 |
| *Rudimentary (i.e., wood, bamboo)* | 6.4 | 0.02-0.13 | 3.3 | 0.01-0.07 |
| *Modern (i.e., zinc, asbestos)* | 85.1 | 0.76-0.92 | 91.9 | 0.87-0.95 |
| Eave open, *n* (%) | 42.6 | 0.32-0.53 | 33.7 | 0.27-0.40 |
| Do windows close, *n* (%) |  |  |  |  |
| *No* | 40.45 | 0.30-0.51 | 25.8 | 0.20-0.32 |
| *Yes – totally closed* | 22.3 | 0.14-0.32 | 20.1 | 0.15-0.26 |
| *Yes – partially closed* | 37.2 | 0.27-0.48 | 54.1 | 0.47-0.61 |
| Vector control measure used, *n* (%) |  |  |  |  |
| *Drains water around the house* | 17 | 0.10-0.26 | 19.6 | 0.14-0.26 |
| *Cuts the grass, keeps lawn clean* | 84 | 0.75-0.91 | 86.6 | 0.81-0.91 |
| *Burns oils or mosquito coils* | 16 | 0.09-0.25 | 14.8 | 0.10-0.20 |
| *Uses insecticides outside of the house* | 7.4 | 0.03-0.15 | 10 | 0.06-0.15 |
| *Nothing* | 6.4 | 0.02-0.13 | 4.3 | 0.02-0.08 |
| **Environment Level** |  |  |  |  |
| Landcover, *n* (%) |  |  |  |  |
| *Woody Savannas (8)* | 37.2 | 0.27-0.48 | 36.4 | 0.30-0.43 |
| *Savannas (9)* | 29.8 | 0.21-0.41 | 39.2 | 0.33-0.46 |
| *Grasslands (10)* | 33 | 0.24-0.43 | 22 | 0.17-0.28 |
| *Croplands (12)* | 0 | 0-0.04 | 2.4 | 0.008-0.6 |
| Elevation (meters), median | 588 | 566-601 | 586 | 575-599 |
| Evapotranspiration at 1-week lag (kg/m<sup>2</sup>/8day), median | 219 | 189-245 | 202 | 174-268 |
| Evapotranspiration at 2-week lag (kg/m<sup>2</sup>/8day), median | 137 | 119-184 | 137 | 95-202 |
| Land Surface Temp Day at 1-week lag (Celsius), median | 35.63 | 35.01-37.4 | 35.27 | 34.87-37.01 |
| Land Surface Temp Day at 2-week lag (Celsius), median | 32.81 | 32.47-32.91 | 32.89 | 32.47-34.43 |
| Land Surface Temp Night at 2-week lag (Celsius), median | 22.73 | 22.57-22.99 | 22.79 | 22.49-22.99 |
| NDVI, median | 0.53 | 0.50-0.54 | 0.51 | 0.49-0.54 |
| Land surface temp night (Celsius), 1- and 2-week lag, median | 22.11 | 21.94-22.19 | 22.13 | 22.05-22.19 |

The most common head of household occupation across malaria status groups was Farmer (52.1% among individuals who had a positive RDT and 38% among individuals that had a negative RDT). The most common highest education level for the head of household among individuals that had a positive RDT result was Primary school 1 and 2 (41.5%), whereas among individuals that had a negative RDT result the most common highest education level for the head of household was Technical School or University (35.4%). Based on the 95%CIs, there is a statistically significant difference between RDT result groups with regard to the percentage of individuals that lived in a house with natural or modern floor material.

### Association between environmental factors and malaria infection

Based on the results of the univariate analyses, the following variables were included as confounders in the final models assessing the association between environmental factors and malaria infection (Table 2): head of household occupation, time of occupation, head of household highest level of education, whether the walls had holes, floor material, whether the windows close, age, and whether they slept under an ITN the previous night. Interactions between the environmental predictors were assessed, however none were statistically significant and thus were not included in the final models. Whether there were holes in the wall was found to be an effect modifier for the relationship between landcover and a positive rapid diagnostic test result and thus was included in the final model for landcover as an interaction term.

**Table 2:**
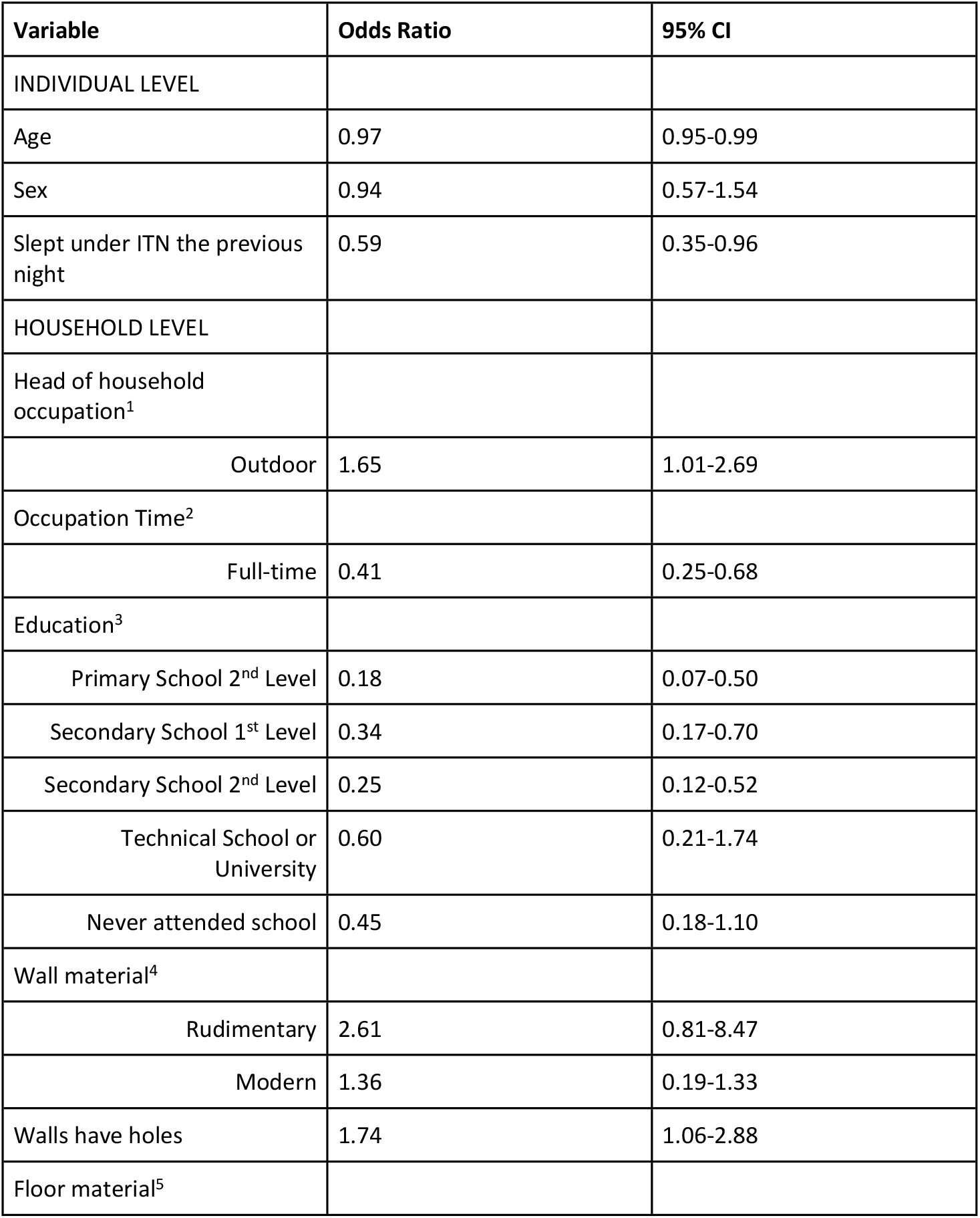
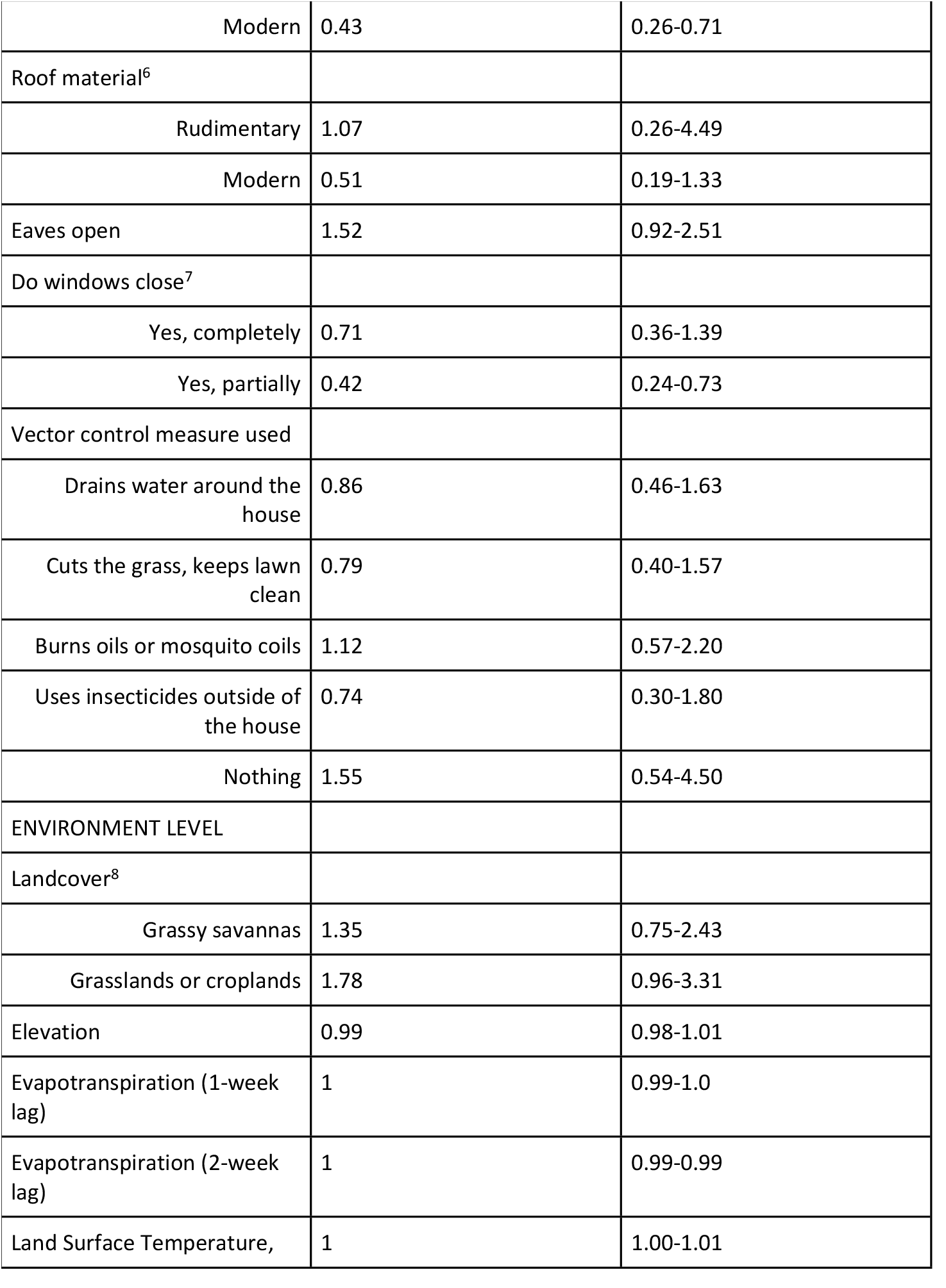
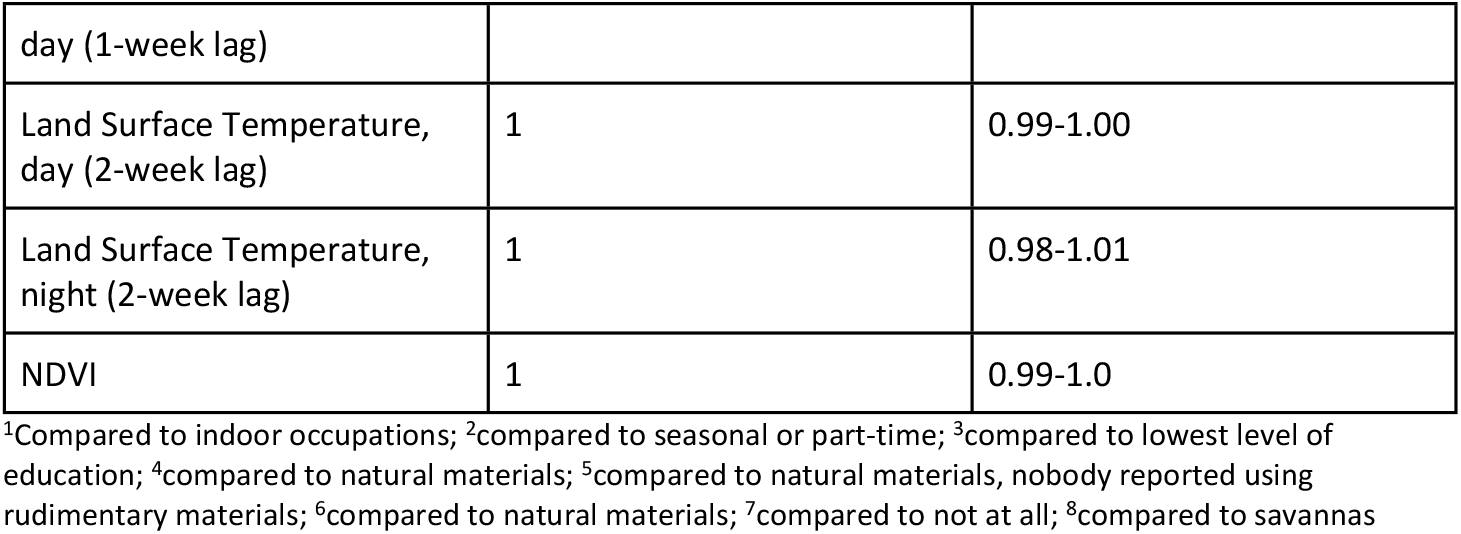
Univariate analyses of individual, household, and environmental level variables

Of the environmental variables assessed, only landcover was found to be strongly associated with having a positive RDT result for malaria (Table 3). Individuals that lived in grassy savannas had 1.55 times the odds of having a positive rapid diagnostic test result compared to individuals who lived in other savannas (OR = 1.55, 95:CI: 0.7-3.7). This result was not statistically significant. The estimated effect of living in grasslands or croplands was found to vary depending on whether the house had holes in the wall (Table 4). Individuals who lived in grasslands or croplands and who had holes in their walls had 5.52 times the odds of having a positive RDT result compared to individuals who did not live in these conditions (OR = 5.52, p-value = 0.035). This result was statistically significant.

**Table 3:** Final environmental models adjusted for individual and household level characteristics

| Variable | Odds Ratio | P-value | 95% CI |
| --- | --- | --- | --- |
| Landcover |  |  |  |
| grassy savannas | 1.55 | 0.316 | 0.7-3.7 |
| grasslands or<br>croplands | 1.27 | 0.568 | 0.6-2.9 |
| Elevation | 0.985 | 0.127 | 0.97-1.0 |
| Evapotranspiration<br>(1-week lag) | 1 | 0.465 | 0.999-1.0 |
| Evapotranspiration<br>(2-week lag) | 1 | 0.085 | 0.9999-1.0 |
| Land Surface<br>Temperature, day (1-week lag) | 1 | 0.058 | 0.9999-1.0 |
| Land Surface<br>Temperature, day (2-week lag) | 1 | 0.318 | 0.995-1.0 |
| Land Surface<br>Temperature, night<br>(2-week lag) | 1.01 | 0.502 | 0.99-1.0 |
| NDVI | 1 | 0.019 | 1.0-1.0 |

**Table 4:**
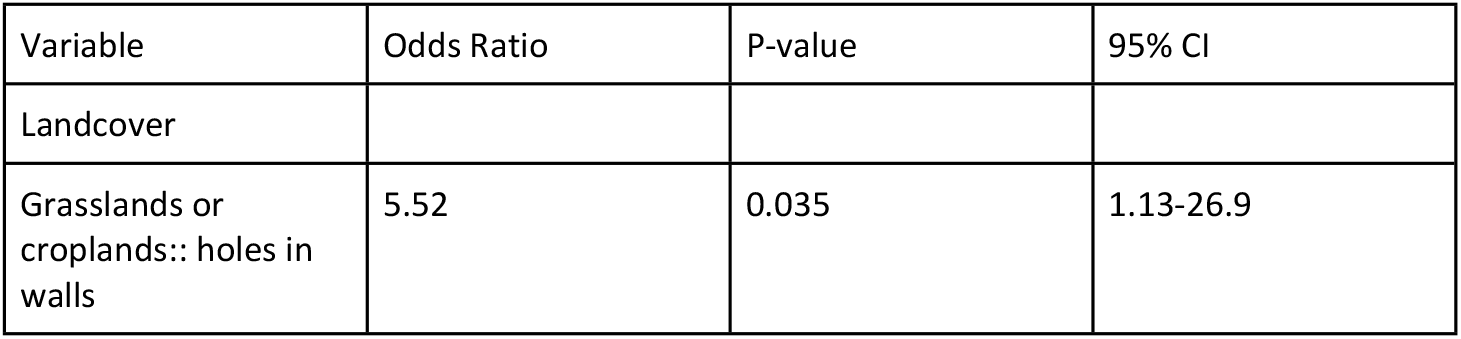
Landcover Model with Holes in Walls Included as Interaction Term

## Discussion

Risk factor surveillance is an important tool for controlling the spread of infectious diseases and in the case of malaria, transmission is greatly influenced by environmental factors. Overall, environmental aspects have not received much attention from malaria control programs. However, understanding how environmental changes impact malaria transmission and infection at the household level could enable local governments to better allocate their resources and optimize disease surveillance and control strategies. We aimed to evaluate the effects of climate and environmental factors on malaria risk at the household level in Sussundenga District, Mozambique. The results from these spatio-temporal models provide information on local characteristics of transmission that can be used to develop frameworks for mitigating malaria transmission that are tailored to this region.

Of the environmental factors that were investigated in this study, landcover was found to be most strongly associated with malaria risk. Moreover, we found that the estimated effect of grassland or cropland landcover on malaria risk varies depending on whether the house has holes in the wall. Thus, housing improvements that could help reduce malaria transmission may include sealing holes in the walls, particularly when the house is located in a grassland or cropland area. This intervention could have a long-term direct benefit that may even go beyond preventing malaria transmission and also affect the transmission of dengue, chikungunya, and other vector-borne diseases. Studies looking at environmental risk factors of malaria infection have found significant associations between infection and temperature and humidity.(9, 10) We did not find an association between malaria infection and evapotranspiration (proxy for humidity) nor land surface temperature day/night at the district level, which may suggest that malaria-environment relationships are contingent on the scale of analysis.

There were some limitations to this study. The MODIS satellite is an optical sensor which means that it cannot observe the surface when cloud cover is present. As a result, there were gaps in the night surface land surface temperature data that we were able to obtain, which is why we ultimately could not assess this variable at a 1-week lag. Additionally, pixels that are partially covered or contaminated by clouds may still have been present in the data which could have affected the study results. This limitation could be addressed in future studies by integrating locally collected environmental data with the satellite data. Lastly, the short time period of the study is a limitation as we could not assess the association between these environmental factors and malaria status across seasons.

## Conclusion

This study reinforces the applicability of the use of remote-sensing data which is available at appropriate spatial and temporal resolution for risk mapping of malaria in areas where weather data is not reliable and readily available. Using remote-sensing data, we assessed the association between key environmental variables and malaria infection at the household level in Sussundenga District, Mozambique. Landcover was found to be most strongly associated with malaria risk, and this association varied depending on whether the house had holes in the wall. These results can be used to develop frameworks for mitigating malaria transmission that are tailored to this region, such as focusing on housing improvements, particularly in grassland and cropland communities.

## Data Availability

De-identified data will be made available upon request to the corresponding author.

## Conflict of Interest

The authors declare that the research was conducted in the absence of any commercial or financial relationships that could be construed as a potential conflict of interest.

## Author Contributions

AS conducted the sourcing of the environmental data, the merging of this data with the epidemiological data, conducted the statistical analyses, and drafted the manuscript. JLF trained and supervised data collection and implementation of the study. ABF, VM, and AN conducted data collection and management as well as community engagement. DEE assisted in the design of study tools, sample selection, and data management. KMS designed the study, implemented the survey used to collect the epidemiological data, and edited and revised the manuscript.

## Funding

This study was funded by a SEED grant from the University of Minnesota Center for Global Health and Social Responsibility.

## Acknowledgements

The authors acknowledge the generous funding from the Center for Global Health and Social Responsibility to complete this study. We also thank the cooperation from the Sussundenga District Health Office, the Manica Provincial Health Office, and the Mozambique Ministry of Health for their collaboration on the study. We also thank the community of Sussundenga village and all study participants for their time and participation in this work.

## Data Availability Statement

Deidentified data will be made available upon request to the corresponding author.

## Notes

### Competing Interest Statement

The authors have declared no competing interest.

### Author Declarations

Ethical review and approval for this study was completed by the Institutional Review Board (IRB) at the University of Minnesota [STUDY00007184] and from A Comissão Nacional de Bioética em Saúde (CNBS) at the Ministry of Health of Mozambique [IRB00002657].

